# Clinical and real-world evaluation of a “fingernail selfie” smartphone app for non-invasive, individually-personalized estimation of blood hemoglobin levels

**DOI:** 10.1101/2022.12.01.22282972

**Authors:** Robert G. Mannino, Parker Lopez, Julie Sullivan, Jeremy Whitson, James Tumlin, Erika A. Tyburski, Wilbur A. Lam

## Abstract

Patients with chronic anemia, or low blood hemoglobin levels, are frequently subjected to the cost, inconvenience, and discomfort of traditional hematology analyzer-based measurements of blood hemoglobin levels via complete blood counts. Elimination of the need for complete blood count testing for hemoglobin screening is an unmet clinical need that we previously addressed by developing a non-invasive smartphone app that estimates hemoglobin levels via image analysis of fingernail bed images. In this work, we present additional data yielding significant improvement upon our previously established technology and describe the clinical validation, and real-world translation of the technology into a commercial product. To improve accuracy and create a clinical use case, we trained the app algorithm on individuals with chronic anemia to personalize the image analysis algorithm for estimating hemoglobin levels. Individual-level differences associated with using the app (variations between individuals, how a user captures images, the specific smartphone they use, the lighting conditions in the location they take the pictures, and biological variability within a population) appear to be the greatest source of measurement variability within larger sample sets. Therefore, we hypothesized that personalization of the algorithm could correct for user-to-user variability and translate to improved accuracy at the individual level.

To test this hypothesis, we trained and tested personalized algorithms for individuals in clinical and “real world” settings. We enrolled 35 chronically anemic subjects [a chronic kidney disease (CKD) cohort] in a clinical study wherein the app algorithm was trained using complete blood count data and paired fingernail bed images, then tested against complete blood count data at subsequent study timepoints. After personalization, testing data revealed a mean absolute error (MAE) of 0.74 g/dL with a root mean squared error (RMSE) of 0.97 g/dL across all testing visits across all subjects, a significant improvement when compared to performance without personalization in the same user group (1.36 g/dL MAE and 1.70 g/dL RMSE, p = 3.13E-11). The app was also used in the “real world” by real app users who self-reported lab/complete blood count blood draw results. App performance findings were consistent with analysis of self-reported data from 17 individuals using our app. After training of the individual app algorithm in the “real world”, testing data revealed a mean absolute error (MAE) of 0.62 g/dL with a root mean squared error (RMSE) of 0.85 g/dL when 4 training data points were used, an improvement when compared to performance of the app without personalization in the same user group (0.71 g/dL MAE and 1.27 g/dL RMSE). ***The personalized app accuracy is similar to that of other noninvasive Hgb measurement technologies currently on the market as medical devices with US Food & Drug Administration (US FDA) clearance. Thus, our technology represents a significant step forward towards true personalized medicine in a digital healthcare setting***.

## Introduction

Anemia, characterized by low hemoglobin (Hgb) levels, affects over 2 billion individuals worldwide each year(1). In the US alone, there are over 83 million people at high risk for anemia and over 10 million people living with chronic anemia, including those with blood disorders, cancer, chronic kidney disease, and nutritional deficiencies(2-5). Those with chronic anemia are routinely screened for anemia via invasive blood tests (complete blood counts, CBCs) that necessitates travel to a clinic, trained technicians, and sophisticated hematology analyzers(6). For some patients, this routine must be repeated multiple times per month.

Smartphones are poised to transform the health and wellness industry. As smartphone use becomes ubiquitous, and smartphone hardware becomes more advanced, powerful computing and imaging technologies are becoming more accessible to global populations (7). Tools to leverage the unique capabilities of smartphones are becoming increasingly available to those suffering from a variety of health conditions, empowering them to live and maintain healthy lifestyles while proactively managing their conditions in collaboration with clinicians(8-10). However, the current clinical paradigm for many health conditions (particularly those of the blood), is centered around expensive, uncomfortable, and time-consuming invasive blood-based testing(6, 11). To advance past these traditional clinical practices, we view the emergence of smartphones as tools that can supplement, enhance, or perhaps even replace existing treatment paradigms, particularly where it comes to invasive testing(12-14). This is especially true in today’s COVID-19 pandemic, where social distancing and over-filled hospitals have limited the availability and accessibility of clinical sites for non-urgent patient visits(15, 16). This is a particularly troubling development for those managing chronic illness and chronic symptoms, including anemia. Anemia management relies on gold standard testing for Hgb level that requires the individual go to a medical facility, undergo an invasive venous blood draw, and wait for the result from the laboratory(17). This paradigm is not only costly and inconvenient, but also a potentially significant cause of health disparities along geographic, age, ethnic, disability, and socio-economic lines(18). With a goal to change this paradigm, we recently reported a noninvasive smartphone app to estimate Hgb levels(19). This app provides users the ability to estimate their Hgb levels noninvasively, from the comfort of their own homes. Access to rapid, Hgb estimates has the potential to enable billions of individuals at high risk for anemia to take more active and proactive roles in their healthcare irrespective of geographic, accessibility, and financial hurdles(20, 21).

The non-invasive smartphone app estimates Hgb levels by quantifying the pallor of fingernail beds via image analysis (Figure 1a). The image analysis algorithm has now been trained on 1,479 user-acquired images and their paired CBC measurements (Figure 1b). Previous clinical assessments determined that the smartphone app is accurate within +/-2.4 g/dL and is suitable for Hgb screening, which can be important in the maintenance of a healthy lifestyle for individuals with typically dysregulated Hgb levels(19). The “fingernail selfie” app was released in the US for both iOS and Android powered smartphones in December 2021 and has 118,533 user accounts and 571,552 hemoglobin estimations completed as of November 30, 2022 at 1:55PM ET (Figure 1b).

**Figure 1:**
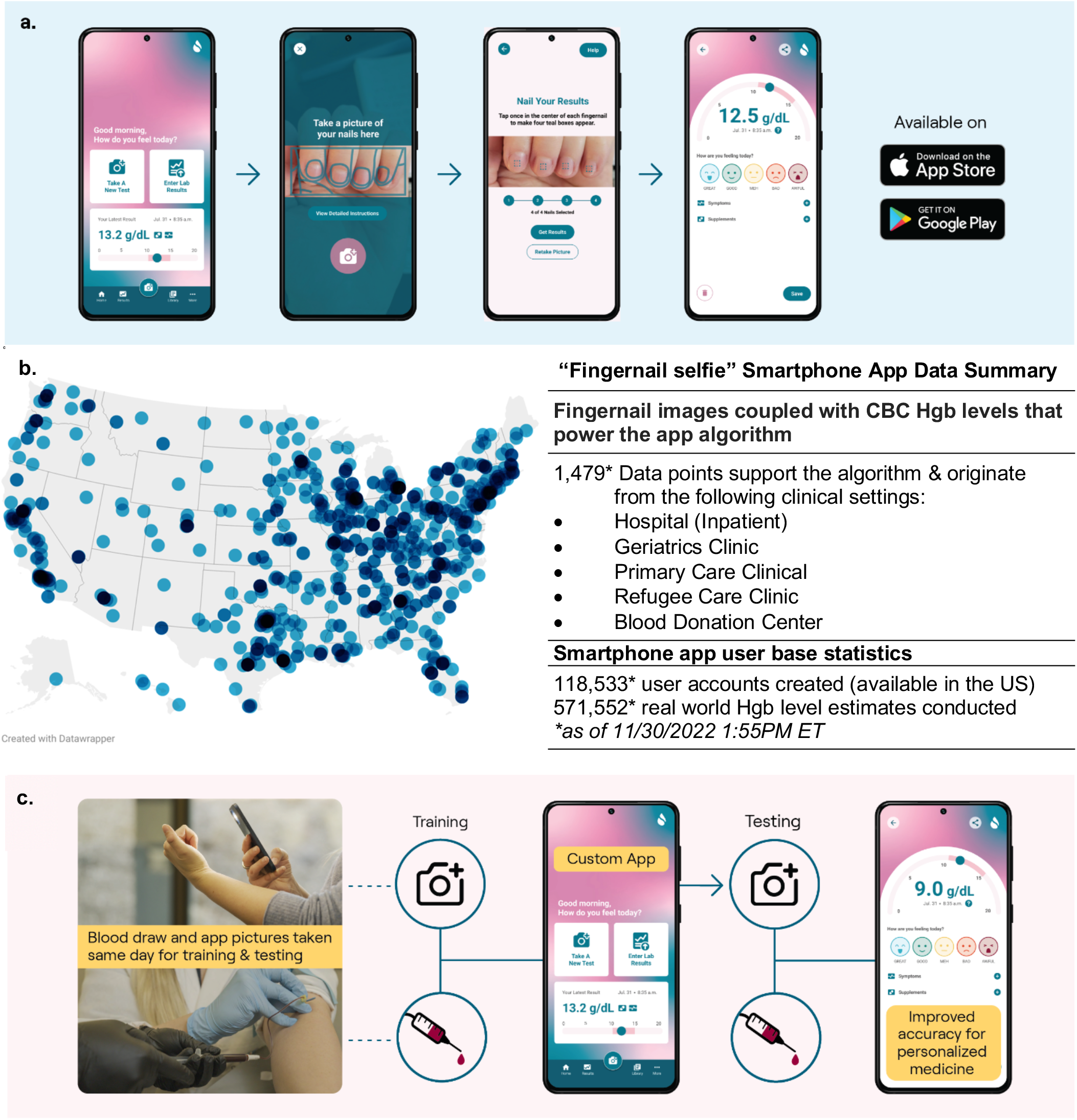
**a. AnemoCheck smartphone app.** The AnemoCheck app works by capturing an image of fingernail beds and calculating how pale the nailbed is. The paleness in the nailbed is correlated to a Hgb level estimate that is displayed on screen. **b. Current smartphone app users and usage**. The smartphone app used in this study is available in the US for both iOS and Android download. Map shows the locations of a random sample of 1,000 smartphone app users across the United States. The table highlights the data breakdown powering the algorithm, and number of app accounts and uses as of November 30, 2022 at 1:55PM ET. **c. Personalized algorithm training and testing**. In this publication, we characterized the effect of training algorithms on the individual level via both a clinically validated study and “real world” data collection. Users captured images of nailbeds on the same days as venous blood draws with blood Hgb levels that were determined by the current clinical standard, a clinical hematology analyzer. The app algorithm was trained by inputting blood Hgb level data with images of nailbeds taken on the same day (training). After the algorithm was trained, the user tested the newly customized algorithm (testing). Results from the customized algorithm were compared with blood Hgb levels taken or reported within 24 hours of app use to establish accuracy of the customized algorithm. Therefore, proof-of-concept was established for improving the app algorithm using personalized calibration in both clinical and real-world examples of personalized medicine.

Whereas our originally described tool is appropriate for Hgb screening and maintenance of a healthy lifestyle, the initial reported accuracy is not sufficient to make diagnoses or inform clinical decisions. However, a more precise and accurate tool could have a significant positive impact on the population of people with known chronic anemia. Individuals who suffer from a variety of conditions such as serious hemoglobinopathies and blood disorders, cancer (and associated chemotherapies), or chronic kidney disease, require clinically accurate means to measure Hgb levels to make diagnoses, mitigate conditions, or inform treatment decisions. We propose that smartphones, enabled with high-performing and validated software, can perform as digital health aids that are personalized to the single individual. Additionally, individual smartphone users can record information pertaining to their specific health condition(s), including symptoms, test results, and medications. This information can then be utilized by digital health tools to personalize those tools and improve accuracy for the individual user. This true example of “personalized medicine” could fundamentally shift the treatment paradigm for a patient suffering from a variety of medical conditions worldwide(22, 23).

In the context of a chronic anemia patient, the ability to personalize a tool to measure their individual Hgb level would enable tracking of Hgb levels without clinical intervention. For example, a transfusion-dependent hemoglobinopathy patient could use this tool to coordinate with their physician to determine exactly when they require their transfusion, as opposed to the guesswork involved with analysis of historical Hgb levels.

Our previous data suggest that most of the error associated with app-based measurements arises from individual variability as opposed to random errors in the image analysis algorithm. Individual variability produces a consistent offset between the app-measured Hgb level and the lab-determined CBC, which we hypothesized could be corrected via individualized algorithm training. To that end, we modified our Hgb estimation smartphone app to permit individual, personalization by taking images using the app on the same day as a CBC/lab blood draw (Figure 1b). This enables the algorithm to be baselined and trained on the individual level, thereby correcting for physiological variability of the finger nailbeds, variability in the way a user may capture an image, smartphone models, and background lighting conditions. This results in user-specific correction factors that are applied to the now-personalized algorithm, thereby improving performance for that specific person. This concept was originally tested in a small pilot on a healthy male, anemic female, and two chronically anemic males, resulting in improvements in the precision and accuracy of the app-obtained Hgb level estimates for individual users(19). Since that preliminary work, we have developed and publicly launched the app as a health and wellness tool for both the iOS and Android mobile environments. **In this work, we present additional data and a major technical advance from our previously reported technology resulting in significant improvement in accuracy, as well as clinical and real-world validation of the tool**.

## Methods and Materials

### Smartphone app and algorithm

Initial app Hgb measurements were done by adapting a previously described algorithm into a smartphone app(19). Hgb calculations are done off-device in a cloud-based infrastructure hosted by Amazon Web Services (Amazon, Inc. Seattle, WA). Cognito (Amazon, Inc. Seattle, WA) is used as our authorization service. Elastic Compute Cloud (EC2, Amazon, Inc. Seattle, WA) instances are used to run the off-device app processes (e.g., calculate the Hgb result, return results history, and manage data traffic and storage) All user data is stored in a MySQL (Oracle, Santa Clara, CA) relational database service (RDS, Amazon, Inc. Seattle, WA), with the exception of user images, which were stored in Simple Storage Service (S3, Amazon, Inc. Seattle, WA), with URL links in the RDS. Locally, the app was loaded onto two smartphone models iPhone 11 Pro [(Apple, Cupertino, CA) running iOS 14 (Apple, Cupertino, CA)] and Samsung Galaxy S20 [(Samsung, Suwon-Si, South Korea) running Android 11 (Google, Mountain View, CA)]. In addition to Hgb estimation capabilities, the app had the ability to create and verify an account, store results history, accept user-entered lab tests, and assign a patient ID.

### Clinical Study

The clinical study characterized the improvement in performance in a chronically anemic patient population suffering from chronic kidney disease (CKD)(2, 24). The clinical team followed patients over the course of eight clinic visits: four “training” visits and four “testing” visits. During each visit, study personnel performed both app-based and CBC Hgb level measurements. During the first four (training) visits, we correlated the app’s estimates with the CBC results to incorporate patient-specific correction factors into the image analysis algorithm. During the second group of four (testing) visits, study personnel used the newly trained app to obtain app Hgb levels and compared them with CBC values obtained via venous blood draw.

### Real World Study

While we were conducting the clinical study, the app was publicly available on both the App Store (Apple, Cupertino, CA) and Play Store (Google, Mountain View, CA). During this period, users were free to download our app and use the same functionality described in this study, namely, the ability to take a test to generate a Hgb estimate, as well as enter in laboratory-determined Hgb levels. Utilizing these self-reported data, we were able to measure the accuracy of the app in a real-world setting, ultimately confirming the results we found in the clinical study.

### Clinical Study Protocol

The study was divided into 2 phases: Personalized algorithm training and personalized algorithm testing.

#### Phase 1 – Training

Clinical study personnel were trained on how to properly capture images with the smartphones according to methods described previously(19). Care was taken to ensure camera flash reflections were avoided and that the fingernails were centered in frame. Following subject screening and written informed consent, study personnel captured images of the subject’s fingernail beds according to instructions provided by the app team. The first four visits were used for training. Immediately after image collection, subjects received a venipuncture blood draw by a trained clinician or lab technician. The resulting blood sample was sent to a 3^rd^ party laboratory (LabCorp – Burlington, NC; Quest Diagnostics – Secaucus, NJ; or PathGroup – Brentwood, TN) for processing on a hematology analyzer, where a complete blood count (CBC) was obtained. When results for the subject were returned, the clinical study coordinator reported the data, along with other demographic and diagnostic information about the subject to the app team. This process was repeated 3 more times for each subject (every 2 weeks +/- 7 days) for a total of 4 training visits that yielded images and paired blood Hgb data points.

#### Intermediate step – App Personalization

In between phase 1 and phase 2 of the study, we analyzed the 4 training images for each subject to create a correction factor specific to each patient (see <u>Data collection and model development</u> section below). This correction factor was then used to update the backend Hgb estimation EC2 instance to automatically adjust the Hgb estimation algorithm according to the subjects’ individual correction factors.

#### Phase 2 – Testing

Subjects completing Phase 1 were invited to continue the study to validate their personalized algorithms. The study allowed for a maximum of 4 testing visits. While using the app, the study coordinator selected a patient’s study ID, which triggered the app to use the subject-specific customized algorithm. The study coordinator then used the app on the subject to determine a Hgb estimate. Immediately after image collection, subjects received a venipuncture blood draw by a trained clinician or lab technician. The blood sample was sent to a 3^rd^ party laboratory (LabCorp – Burlington, NC; Quest Diagnostics – Secaucus, NJ; or PathGroup – Brentwood, TN) for processing on a hematology analyzer, where a CBC was obtained. When results for the subject were returned, the clinical study coordinator recorded the data, along with other demographic and diagnostic information about the subject to the app team. This process was repeated a maximum of 3 more times for each subject (every 2 weeks +/- 7 days) with between 1-4 testing images and comparison blood Hgb data points being collected. This comparison was used to validate each subject’s personalized correction factor. After the final test, the study coordinator conducted a closeout survey to complete the study. Subjects were compensated $100 per visit for the duration of the study.

In total, 40 subjects were enrolled in this study. Of that cohort, 35 subjects completed phase 1 of the study and had at least 1 phase 2 visit. Both phases were completed by 32 subjects. One subject of those 32 was removed from the study due to fingernail discolorations that were determined to meet the exclusion criteria after enrollment and data collection. In the original cohort of 40 subjects, two subjects unfortunately passed away before completion of the study due to their underlying conditions, and 6 subjects voluntarily withdrew from the study at various stages.

### Data collection and model development

The data collection process began with the capturing of images via the smartphone application in phase 1. The clinical coordinator managed this process and notified the study coordinator when each subject visit occurred to alert the team to new images and data. Following notification, the study coordinator coordinated with our data science team to perform quality assurance on each image. The study coordinator then notified the clinical coordinator of any potential issues in the data. After passing QA, the data was added to a master study enrollment log containing all deidentified patient data. The photos from the subject visit were then added to an image database for analysis.

Model adjustment parameters were built for each patient using their phase 1 data. Calculation of each patient’s personal adjustment parameter was completed by first converting the data for that patient into a .csv file. A script was then used to extract all the necessary data required for Hgb measurement(19), which was output a Pandas data frame or Python dictionary depending on use. After this processing step, data from three replicate images for each phone model at each patient visit were averaged together, and from that an offset was computed relative to the base Hgb estimation model. This relative offset was computed for each patient visit and the average of the four training visits was calculated. The resulting average was a patient-specific correction for each smartphone device. These parameters were then recorded and used to update the smartphone application source code to implement the subject specific models. For every subsequent visit after the fourth visit (phase 2 testing), the corrected model was used to approximate the Hgb levels for a patient.

### Data privacy and Safety

#### Clinical data

All patient data were de-identified by the clinical team prior to sharing with the research team. No personally identifiable data were shared with the research team. All protocols and procedures for the clinical study were approved by an institutional review board (Advarra IRB, Pro00049129).

#### Subject population

The goal of this study was to enroll 40 patients with CKD within stage 3-5 to represent a chronically anemic population. The study enrolled 40 patients with CKD of various stages (20 Stage 3, 16 Stage 4, 4 Stage 5). Eighteen patients had diabetes. Of this total, 6 subjects voluntarily withdrew from the study prior to completion and 2 subjects passed away prior to completing the study due to health complications unrelated to the study. The following exclusion criteria were used to determine enrollment:

- Subjects with chronic persistent esophageal reflux disease unless being actively treated for it;
- Patients with known Gastric or Duodenal peptic ulcer disease during the previous 6 months;
- Subjects undergoing chemotherapy for oncologic or auto-immune disorders;
- Subjects who were wearing, or within 30 days of enrollment had worn, artificial nail products (SNS, acrylic nails etc.);
- Subjects who were wearing traditional nail polish and were unable or unwilling to remove nail polish;
- Subjects with nailbed obstructions or discolorations (e.g., leukonychia, melanonychia, bruising, etc.).

### Real World (Self-Reported data)

#### Hgb Screening

The currently available smartphone app has a feature that allows users to enter laboratory results for tracking purposes. We analyzed real-world user data from our app and filtered the data set down to the 765 times that a user entered a laboratory Hgb level into the app and received a Hgb level estimate using the app within 2 days of that lab Hgb result. Each lab result was compared to its closest app result (within 2 days) and accuracy statistics were calculated based on these comparisons. Reported app results below 7 g/dL and above 17 g/dL were excluded from the study since the underlying algorithm was not trained on data outside that range and thus was not designed to evaluate Hgb levels outside of that range. Furthermore, this was done to perform quality control on the entered laboratory values (e.g., we received 19 entered lab results below 1 g/dL, which is highly improbable biologically, and likely was the result of user error when manually entering the data). A cutoff of 2 days between app test and lab test was chosen to minimize the likelihood of Hgb levels shifting between the lab test and app test, which can occur over the timescale of 2 days. Furthermore, this timeline helps ensure that the user has an accurate memory of the laboratory test when self-reporting.

#### App Personalization

Of the 538 users who entered their own lab tests, 16 utilized this functionality 5 or more times. This allowed us to replicate the methods of our clinical study to develop a personalized algorithm correction for those users. We treated the first 4 entered lab tests as the “training” phase, and the results that followed result 4 as the “testing” phase. A total of 64 training points and 40 testing points were acquired across all subjects.

### Statistical analysis

Paired t-tests for means were used to determine the statistical significance of the improvement of app accuracy when calibrated using both real world and clinical study data. Two-sample t-tests were used to determine the statistical significance of the difference in app accuracy when analyzed by cohort.

## Results

### App Training improves Hgb estimation

Personalized training of the Hgb estimation model led to significant (p = 3.13E-11) improvements in accuracy (Figure 2). Personalized corrections were developed using data from phase 1 of the study (half the study involvement). A total of 35 chronically anemic subjects (a chronic kidney disease subject cohort) completed the pilot study (phase 1: 4 study visits to collect training data and phase 2: between 1-4 study visits per subject to collect testing data). Testing data revealed that the MAE was 0.74 g/dL with a root mean squared error (RMSE) of 0.97 g/dL (n = 34 subjects) across all testing visits across all subjects, compared to 1.36 g/dL and 1.70 g/dL, respectively, when untrained (Figure 2a & 2b). This improved performance shows that the RSME for the personalized smartphone app is lower than that observed for FDA-cleared non-invasive Hgb determination methods (RSME of 1.1), suggesting superior performance over those clinically validated tools(25).

**Figure 2:**
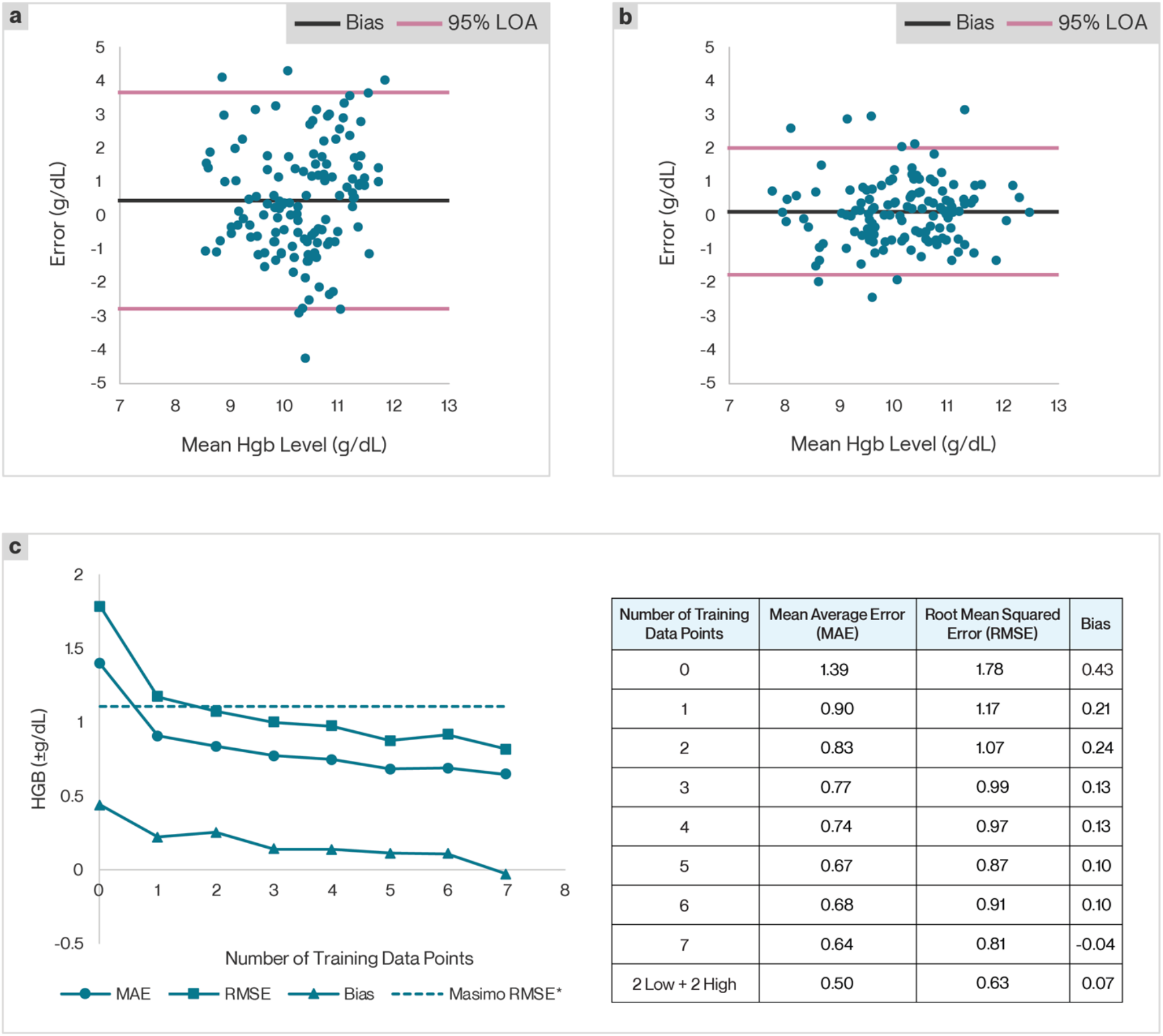
Performance improves with customized trained app. A total of 40 subjects were enrolled in this study. Of that cohort, 35 subjects completed phase 1 of the study and had at least one phase 2 visit. 32 subjects completed both phase 1 and phase 2 and one subject was excluded. In the first four visits (Phase 1), images of fingernail beds were captured along with a venous blood draw for blood Hgb level. **a**. First, these data were compared against blood Hgb levels to serve as a control for app results without app training (n=35). Then, these data were used to train a custom algorithm for each subject. **b**. In visits five through eight of the study, the personalized algorithm was tested and compared with the results of venous blood draws taken during those visits (n=35). The average error of the personalized algorithm testing data points (visits 5-8) was 0.74 g/dL with a root mean square error of 0.97. Bias was 0.16 g/dL, which represented an improvement over the untrained app (n=35). **c**. App performance improves with increasing number of training data points (n=31). Subject data across 8 data points was analyzed assuming different numbers of training points and testing points. Average error, root mean squared error, and bias of the testing points are shown for the number of training data points. We have also included a data point in the table using the two lowest and two highest blood Hgb level data points to train the algorithm. The results indicate that the average error, root mean squared error and bias are all smaller when the algorithm is trained with data across a broader Hgb level range.

### Increasing the number of training points improves Hgb estimation

We found that the number of training points directly correlated with improved accuracy of the Hgb estimation model (Figure 2c). As we added training points, Hgb estimation improved from a mean absolute (MAE) of 1.39 g/dL and RMSE of 1.78 g/dL to a MAE of 0.64 g/dL and RMSE of 0.81 g/dL with 7 training points (n = 31 subjects). **Importantly, RMSE drops below 1.1 g/dL after only 2 training points, a threshold established by the United States Food and Drug Administration as sufficient for clearance as a class II medical device for noninvasive hemoglobinometers(25)**.

### Comorbidities such as CKD severity and diabetes have little impact on Hgb estimation

Finally, we show that diabetes and CKD, two major comorbidities with and causes of anemia, have little impact on Hgb estimation. In this group of subjects, those without diabetes had a statistically negligible improved accuracy compared to those with diabetes (MAE 0.77 g/dL and 0.71 g/dL respectively, p = 0.27). When analyzing Hgb estimation accuracy by severity of CKD, we again found a statistically negligible decreased accuracy as CKD progressed from stage 3 to stage 5 (Stage 3 to Stage 4 - MAE = 0.66 g/dL, 0.83 g/dL respectively, p = 0.06. Stage 4 to Stage 5 - MAE = 0.83 g/dL, 0.75 g/dL respectively, p = 0.33. Stage 3 to Stage 5 - MAE = 0.66 g/dL, 0.75 g/dL respectively, p = 0.23) (Table 2).

**Table 2:** Comorbidity impact on Hgb estimation.

|  | CKD Stage | Mean Average Error | RMSE |
| --- | --- | --- | --- |
| 3 | 20 | 0.66 | 0.86 |
| 4 | 16 | 0.83 | 1.06 |
| 5 | 4 | 0.75 | 1.06 |

|  | Diabetes | Mean Average Error | RMSE |
| --- | --- | --- | --- |
| Yes | 18 | 0.77 | 1.00 |
| No | 22 | 0.71 | 0.93 |

### App performance translates outside of the clinic

Furthermore, we analyzed self-reported data on 16 users who reported 5 or more laboratory tests with app estimates within 2 days and applied this personalized algorithm approach to these subjects (Figure 3). Personalization of the app improved performance, from an MAE of and 0.71 and a RMSE of 1.27 when uncalibrated (Figure 3a), to an MAE of 0.62 g/dL and an RMSE of 0.85 g/dL following calibration (Figure 3b). This increase in accuracy follows the trend reported in the clinical study data and increasing the number of training points improves accuracy (Figure 3c). In our previous publication, we described the performance of the image analysis algorithm for correlating nailbed pallor to blood Hgb levels, as well as the accuracy of Hgb level estimation (Mean Absolute Error (MAE) = ±1.0 g/dL, 95% LOA = ±2.4 g/dL)(19). In this work, we analyzed Hgb estimates recorded by the app in our publicly available wellness tool compared with self-reported Hgb levels and report similar performance in Figure 3d. In individuals with Hgb > 10g/dL, the intended target of our app, MAE = ±0.6 g/dL, RMSE = ±1.1 g/gL, 95% LOA = ± 2.1 g/dL (n = 673 measurements from 474 individuals)

**Figure 3:**
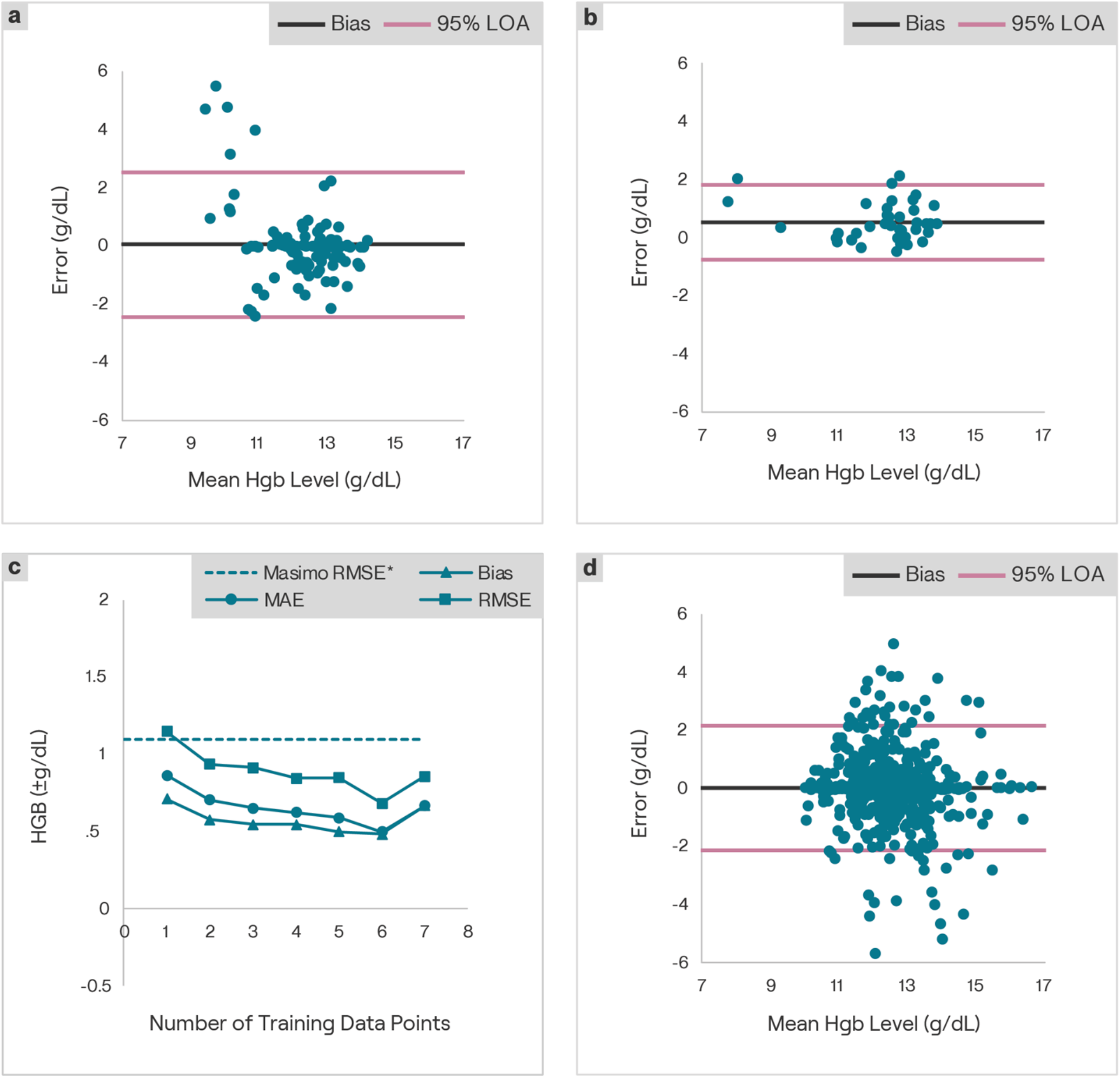
Real world use of the app validates clinical findings. Real world app users and their self-reported data were used to complete app training and testing. **a**. First, 104 data points from 16 users who self-reported at least 5 lab tests were used to serve as a control for app results without app training. Comparing these data to the entered lab-derived Hgb levels resulted in a mean absolute error of the personalized algorithm testing data points was 0.71 g/dL with a root mean square error of 1.27 g/dL. Then, the 64 data points corresponding to each user first 4 entered laboratory Hgb results were used to train a custom algorithm for each user. **b**. The newly customized algorithm was tested with the next 40 data points and compared with the results of self-reported blood Hgb levels reported within 24 h of using the app. The mean absolute error of the personalized algorithm testing data points was 0.62 g/dL with a root mean square error of 0.85 g/dL, which represented an improvement over the untrained app in the “real world”. **c**. App performance improves with increasing number of training data points. Real world subject data across 8 data points was analyzed assuming different numbers of training points and testing points. Average error, root mean squared error and bias of the testing points are shown for the number of training data points. **d**. In total, 474 users entered 673 lab tests with Hgb levels > 10 g/dL. In these users, the mean absolute error and RMSE compared to the entered lab results was 0.61 g/dL and 1.1 g/dL, respectively.

## Discussion

The previously described uncalibrated algorithm in the app is intended to be used for Hgb estimation and screening in the general population and maintenance of a healthy lifestyle. In this work, we confirm the accuracy of the healthy lifestyle tool based on self-reported data in the real world. This is an important result as it displays the versatility of the app technology to adapt to numerous introduced sources of error that exist when an experimental technology translates from the bench to the bedside. In our previous clinical study, results were reported using a single smartphone, with one trained user, in a very consistent imaging location (various rooms within an academic medical center), using a single gold standard Hgb reference. The current algorithm has been trained on 1,479 data points from subjects in the United States as of November 30, 2022 at 1:55PM ET. Since the previously described app publication, we have released the app publicly as a wellness tool in the US and analyzed data from hundreds of users with different smartphones that self-reported their Hgb levels, illustrating the truly translational and personalized nature of our technology.

In this work, we take the technology one step further and show that the app can be calibrated for individual users in a chronically anemic population. The accuracy of the smartphone app when compared to a blood test Hgb level is significantly improved by calibrating the app. The primary reason for this significant accuracy improvement lies in the ability to compensate for several variables that are very difficult to control for in the real world. App personalization leverages the user as their own control and controls for individual biological user variability, as well as for the environmental conditions such as background lighting, smartphone model, and user experience (i.e., the unique way individuals interact with a tool). Furthermore, the consistent results between the clinical study on CKD patients and the self-reported study indicates that users are willing and able to perform the calibrations necessary, since hundreds of users utilized the “enter lab test” functionality when personalization was not even an advertised feature of the app; users were unaware of the potential app personalization benefits associated with blood test result entry. Personalization improved the Hgb estimation performance of the app, yielding an accuracy that is comparable to other noninvasive Hgb determination devices that have been cleared by the US FDA for clinical Hgb level measurement(25). ***Overall, the improvement in Hgb estimation accuracy [decreases in root mean squared error (RMSE) and decrease in absolute difference between CBC blood Hgb level and app Hgb level] brings the performance of this technology in line with commercially available, FDA-cleared devices for Hgb level determination, significantly enhancing the clinical utility of this technology to aid in diagnosis and treatment of anemia. The self-reported data we collected not only supports this contention, but it demonstrates the desire of individuals around the country to use a tool like this and their ability to successfully do so***.

It is also important to note that it only takes 2 training points to bring the accuracy to comparable levels to US FDA-cleared methods, with further performance improvements associated with the addition of more training points. This provides significant flexibility for the user and their healthcare provider. This flexibility has the potential to accommodate many different anemia-causing diseases and conditions with different treatment regimens requiring blood draws. This flexibility allows users and their physicians to balance the inconvenience of blood draws for app training with accuracy gains derived from them. For example, conditions that require less frequent Hgb monitoring can still benefit from a trained app that reduces the need for expensive clinical methods. However, if a user’s condition necessitates frequent clinical visits and blood draws, they can utilize this to their advantage and make significantly improved app just by following their existing blood testing schedule. Overall, the findings in this pilot study justify a larger multi-site study to test the personalized technology on additional cohorts of subjects with chronic anemia against gold standards and other non-invasive methods to confirm clinical utility.

Furthermore, real world testing confirms the translational capability of this technology. As of preparation of this manuscript, the publicly available app has been used by over 100,000 people in the United States. Using self-reported laboratory-determined Hgb levels from hundreds of users, we were able to confirm the results of our clinical testing, a result made possible by the widespread accessibility of this system. This smartphone application is the first of its kind to enable truly personalized medicine, where patients with chronic anemia can tailor a completely noninvasive and instantaneous smartphone app to measure and track Hgb levels, with an accuracy comparable to clinical methods.

Key limitations of the clinical study include the consistency of the imaging parameters, and the small sample size for comorbidity analysis. The clinical study took place at 2 nearly identical clinical practices. Images were taken on only 2 smartphone models by study personnel. This situation negates much of the variability seen in at-home testing environments (e.g., different room lighting, user error, device heterogeneity, etc.), the intended environment for this app. While the variability introduced by these environments has not been tested to date, they can be mitigated. We have previously reported that use of the camera flash overpowers background lighting conditions to normalize the background brightness of an image(19). Furthermore, the use of a consistent location, user, and phone model for training allows for each of these variables to be controlled. A user could select a single location (this could even be the clinic setting they receive their blood draws in – likely closely matching our study parameters), using their personal smartphone, allowing the training phase to take all these variables into account. Key limitations of the real world self-reported data are the inability to control or verify that the app was used correctly or that the entered lab tests are accurate. However, we would expect significantly discordant results if these sources of error were impacting app performance. The fact that self-reported results were consistent with clinical study results indicates that the impact of these potential errors outside of the clinic is minimal.

The presence of comorbidities with anemia presents a double-edged sword. We show that we can tailor our Hgb estimation models for specific diseases, however, the potential variability introduced into the system by the comorbidities must be addressed. Our limited comorbidity analysis showed trends of worsening app performance with worsening disease (i.e., Hgb estimation was less accurate with diabetes than without, and Hgb estimation accuracy got worse as CKD severity progressed), however, the observed effect was not great enough to produce statistically significant differences. Ultimately, the findings of this limited study support the conclusion that neither diabetes nor CKD progression had significant impacts on Hgb estimation accuracy and supports our rationale for using fingernail beds as image subjects. As the Hgb in the blood vessels within the nailbed are the primary pigment contributor to nailbed color, the cause of anemia should not matter, however, further study is needed to confirm the impacts that other diseases have on app performance, particularly etiologies that impact fingernail color outside of Hgb (e.g., jaundice, cyanosis, poor circulation, etc.)(26-28).

Overall, we have continued to improve upon our original concept for noninvasive measurement of Hgb levels using only a smartphone, which could enable billions of individuals of users worldwide to track their Hgb levels from the comfort of their own homes. Personalization via training of our non-invasive smartphone application for Hgb level determination improves the accuracy to that of other US FDA-cleared non-invasive Hgb determination methods and findings justify a larger multi-site study to test the personalized technology on additional cohorts of subjects with chronic anemia against gold standards and other non-invasive methods to validate clinical utility. Results from this work support the rationale that patient-obtained clinical results can be incorporated into digital health tools to enhance their performance by customizing the functionality of those tools. Approaches such as these offer the promise of bringing true “personalized medicine” to fruition. The combination of app, methods and results presented here is the first of its kind to bring personalized medicine to the realm of blood testing. Furthermore, real world use of the app directly illustrates the translational potential of this technology, as we have a large userbase currently utilizing the app as well as the functionality that enables this personalization. This app has the potential to equip patients and their physicians with a tool for managing treatment and care beyond the clinic and empowers patients to play a more active role in their health, with a medical tool that is truly unique to them. Ultimately, the work presented here has the potential to transform the digital health landscape and serve as a model for deploying smartphones as clinical tools in personalized medicine.

## Data Availability

All data produced in the present study are available upon reasonable request to the authors.

## Acknowledgements

Funding for this study was provided in part by the National Heart Lung, and Blood Institute (NHLBI) of the National Institutes of Health (NIH) via award 2R44HL139250-02A1 as well as by AstraZeneca.

## Declaration of Interests

R.G.M, P.L, J.S, E.A.T, and W.A.L are all employed by Sanguina, Inc and have an interest in the technology described in this study. Furthermore, R.G.M, P.L, E.A.T, and W.A.L are equity holders in Sanguina, Inc.

## Notes

### Author Declarations

The Advarra institutional review board gave ethical approval for the clinical work (Pro00049129).

